# Microscopic fractional anisotropy MRI differences in genetic frontotemporal dementia

**DOI:** 10.64898/2026.05.25.26354046

**Authors:** Isis So, Ricardo Rios-Carrillo, Kristy K. L. Coleman, Elizabeth C. Finger, Corey A. Baron

## Abstract

**INTRODUCTION:** Microscopic fractional anisotropy (µFA), an emerging diffusion MRI metric, may be more sensitive than conventional metrics to gray matter microstructural changes in neurodegeneration. This pilot study compared µFA, mean diffusivity (MD), and volume between genetic frontotemporal dementia (FTD) variant carriers and non-carriers in the insula, frontal pole, and medial orbitofrontal cortex (mOFC).

**METHODS:** Carriers and familial non-carriers of FTD variants in *C9orf72, GRN*, or *MAPT* were scanned between October 2024-December 2025. Non-parametric aligned rank transform ANCOVAs were computed to analyze between-group differences in µFA, MD, and volume while controlling for age.

**RESULTS:** Carriers (*n*=12) exhibited lower insula µFA than non-carriers (*n*=8): *F*(1,19)=5.89, 95% CI [-10.7,-0.75], *p*=0.027, *η*^*2*^*p*=0.26. No group-differences were observed in other metrics, including MD and volume.

**DISCUSSION:** Reduced µFA in the insula, a region vulnerable to early atrophy in FTD, may be more sensitive to early microstructural changes in genetic FTD than traditional diffusivity measures.

## INTRODUCTION

Frontotemporal dementia (FTD) is a rare and early-onset neurodegenerative dementia characterized by impairments to behaviour, memory, and language.^1^ Approximately 10-25% of genetic FTD cases are caused by autosomal dominant variants in one of three genes: chromosome 9 open reading frame 72 (*C9orf72*), microtubule-associated protein tau (*MAPT*), or granulin (*GRN*).^1^ The average age of onset differs per form of genetic FTD and can vary widely, with *C9orf72*-associated FTD at ∼58 years (range: 20-91), *MAPT*-associated FTD at ∼49 years (range: 25-90), and *GRN*-associated FTD at ∼61 years (range: 17-82).^2^ However, studies from large-scale international FTD cohorts have revealed differences in brain volumes,^3–9^ structural integrity,^10–13^ and functional connectivity^7,14,15^ detectable on MRI in presymptomatic individuals in the years and decades prior to symptom onset.

Volumetric MRI studies which compared presymptomatic FTD variant carriers (collapsed across *C9orf72, MAPT*, and *GRN*) and non-variant carriers detected reduced volumes in the insula and temporal lobe approximately 10 years prior to symptom onset in carriers, and reduced volumes in the frontal lobe, amygdala, striatum, and thalamus around 5 years prior to disease onset.^3^ A mix of cross-sectional^4–7^ and longitudinal analyses, and modelling based on estimated years to disease onset,^8,9^ have also revealed regional volume losses specific to *C9orf72, MAPT*, and *GRN* which can emerge up to two to three decades before symptoms occur. Diffusion MRI findings have likewise revealed white matter integrity losses, via both reduced fractional anisotropy (FA) and increased mean diffusivity (MD), during presymptomatic stages of genetic FTD in the uncinate fasciculus, corpus callosum, cingulum, and corticospinal tracts in *C9orf72* carriers, and the uncinate fasciculus in *GRN* and *MAPT* carriers.^10^ Gray matter integrity loss, indicated by increased MD, has also been observed in thalamic subnuclei of small samples of presymptomatic carriers of all three genetic FTD subtypes (median ages of 37.1, 43.5, and 36.0 years for *C9orf72, GRN*, and *MAPT*, respectively),^13^ and in the hippocampus and amygdala of asymptomatic *MAPT* carriers (average age of 36 years).^12^

While replicated findings support that volume and microstructural losses occur in presymptomatic carriers, remaining questions include whether earlier or larger differences can be detected by more sensitive non-invasive neuroimaging methods, and what potential mechanisms underlie such early changes. Recent advances in diffusion MRI can enable a more granular characterization of MRI differences. This includes the application of biophysical models of diffusion MRI in FTD, such as neurite orientation dispersion and density imaging (NODDI), ^11,12^ which enable characterizing neurite microstructure with greater specificity. Combining NODDI with DTI measures in a machine learning algorithm can be more accurate than conventional DTI measures alone in classifying FTD subtypes.^11^ Some NODDI metrics, including the orientation and dispersion index, can also be more sensitive in temporal lobe regions and long association and projection fibres in young asymptomatic *MAPT* carriers (average age: 36 years) compared to conventional DTI metrics, including MD.^12^

While NODDI has been relatively well-validated in white matter, the biophysical assumptions made by this model have weak compatibility with grey matter. A relatively new acquisition approach for diffusion MRI enables computation of microscopic FA (µFA), which is sensitive to diffusion anisotropy regardless of relative fibre orientations within a voxel, and allows a measure of fibre content not confounded by crossing fibres. These properties are advantageous over conventional FA, which is affected by both axonal microstructure and fibre orientation. Furthermore, given that µFA requires no microstructural assumptions, it can be applied to white or grey matter without the caveats of microstructural modelling.^16^ µFA thus potentially enables a more granular understanding of both white and gray matter regions with complex fibre architecture, including regions which represent structural hubs, and may be more sensitive to fibre loss in regions with crossing fibres compared to FA or NODDI metrics.^17^ µFA has been shown to be more sensitive than conventional diffusion MRI metrics for detecting age-related microstructural changes in healthy individuals,^18^ cognitive training-induced white matter changes,^19^ hippocampal sclerosis in drug-resistant temporal lobe epilepsy,^20^ lesions in multiple sclerosis,^21^ and white matter degeneration in Parkinson’s disease.^22^

Applying this novel diffusion MRI metric to genetic FTD for the first time, this pilot study examined µFA, MD, and volumes in genetic FTD variant carriers compared to non-carriers in three hypothesis-driven regions of interest (ROIs): the insula, frontal pole, and medial orbitofrontal cortex (mOFC). These ROIs were chosen because volume and microstructural differences are consistently detected earliest in these regions when *C9orf72*-, *MAPT*-, and *GRN*-associated FTD are examined together.^3,4,8,9^ It was posited that µFA of these ROIs would be reduced in carriers compared to non-carriers, and that µFA group differences would be larger than those detected using MD or volumetry.

## METHODS

### Participants and Study Design

This pilot observational cohort study recruited 22 adults between October 2024 to December 2025 who were carriers or familial non-carriers of FTD pathogenic variants in *C9orf72* (>30 repeats), *GRN*, or *MAPT*. Participants underwent T1- and a diffusion-weighted MRI protocol optimized for rapid µFA measurements.^23^ An exclusion criterion included participants with unknown genetic carrier status, which resulted in *n*=20. Cross-sectional µFA measures were compared between FTD variant carriers and familial non-carriers.

Ethics approval was obtained at the local institutional research ethics board. All participants provided written informed consent, in line with the Declaration of Helsinki.

### Neuroimaging

Participants underwent T1- and T2-weighted MRI based on the GENFI3 protocol using a 3T Siemens Prisma scanner. Both anatomical scans had the following sequence parameters: FOV=256 × 256mm^2^, 208 slices, 1.1mm isotropic voxel size, and 8° flip angle; the T1-weighted anatomical scan was a Magnetization Prepared Rapid Gradient Echo (MPRAGE) image with TR/TE=2000/2.85ms and TI=850ms; the T2-weighted anatomical scan had TR/TE=3200/401ms.

Participants then underwent a diffusion MRI protocol to estimate µFA measurements. The Siemens diffusion echo-planar imaging (EPI) sequence was used, with diffusion gradients modified for spherical tensor encoding (STE; required for µFA computation) in-house.^23^ The parameters included: TR/TE=5000/92ms, simultaneous multislice (SMS) factor 2, rate 2 generalized autocalibrating partially parallel acquisitions (GRAPPA) acceleration factor 2, FOV=229 × 229 mm^2^, and 1.8mm isotropic voxel size. The protocol included 8 linear tensor-encoded volumes at b=2000s/mm^2^ and STE acquisitions consisting of 3, 6, and 16 volumes at b-values of 100, 1000, and 2000s/mm^2^, respectively. Total acquisition time was 3:09 minutes. This protocol was computed twice using opposite phase-encoding directions: anterior-to-posterior, followed by posterior-to-anterior.

Diffusion MRI data were preprocessed using principal component analysis denoising and Gibbs’ ringing artifact correction via *dwidenoise*^24^ and *mrdegibbs*^25^ in MRtrix v3.0.4.^26^ Subsequent correction for EPI readout distortions and eddy current effects was performed using *topup*^27^ and *eddy*^28^ from FSL v6.0.7.17.^29^ T1-weighted images underwent cortical and subcortical segmentation using v7.4.1 of FreeSurfer’s probabilistic atlas-based parcellation^30^ to obtain the ROIs. T1-weighted images were registered separately to the 1.8 mm diffusion MRI spaces using linear affine registration with a mutual information similarity metric implemented in Greedy v1.3.0 (https://greedy.readthedocs.io/en/latest/),^31^ and MD and µFA maps were calculated using MatMRI (https://gitlab.com/cfmm/matlab/matmri).^32^ The resulting inverse transformations were applied to register MD and μFA images into anatomical space. Registration quality for all scans was assessed by visual inspection.

### Statistical Analysis

All analyses were computed using R v4.4.1. Differences in age between carriers and non-carriers were determined using a t-test. Data distribution was visualized using histograms and q-q plots, and numerically defined using the Shapiro-Wilk test. While the residuals of the dependent variable of interest, µFA, did not violate data normality assumptions, sample sizes were small, which may limit the robustness of parametric inference and the reliability of variance estimates. A non-parametric aligned rank transform ANCOVA, which requires an ordinal dependent variable, was thus chosen to provide more robust statistical inference while retaining the factorial model structure. The assumptions for an aligned rank transform ANCOVA, including independence of observations, and ranks preserving meaningful ordering of the outcome variable, were met.

Participants were grouped by carrier status combining *C9orf72, MAPT*, or *GRN*, and analyses were not segregated further by each genetic group due to sample size. One model was computed per ROI for the primary outcome, µFA, where each model compared rank aligned µFA between groups while controlling for age. To assess whether µFA was a comparable MRI marker to conventional volumetric and diffusion MRI metrics, models per ROI were also computed for volume and MD while adjusting for age. Since MD can be more sensitive than FA for detecting early white matter microstructure changes in FTD,^15^ we chose to compare MD instead of FA. Effect sizes were examined with partial eta-squared (*η*^*2*^*p*). Statistical significance was defined as *p*<0.05. Given the hypothesis-driven determination of the limited number of ROIs, correction for multiple comparisons was not performed.

For any ROI with µFA differences, a post-hoc model was computed to determine whether µFA effects were more pronounced in symptomatic patients. This model included symptomatic status as the independent variable (levels: symptomatic carrier, pre-symptomatic carrier, or non-carrier as the reference) while controlling for age.

## RESULTS

Demographic comparisons revealed that FTD variant carriers (*n*=12; *n*=7 presymptomatic; *n*=5 symptomatic, with average 5.2 years since diagnosis) and familial non-carriers (*n*=8) did not differ in age (*t*=-0.69, *p*=0.50, *d*=-0.31) **(Table 1)**.

**Table 1.**
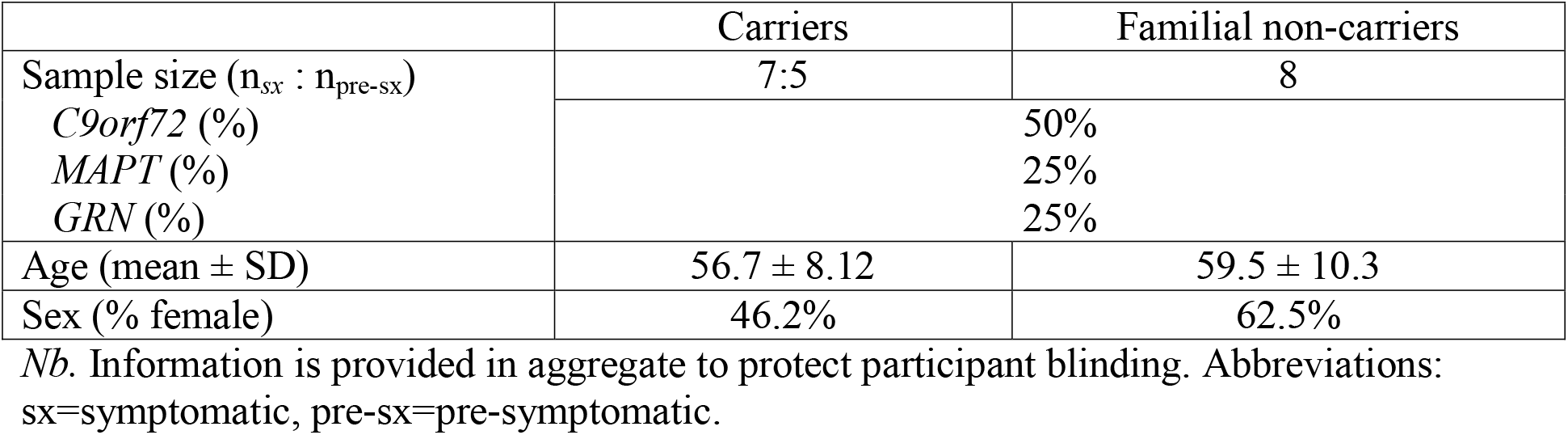
Demographic characteristics of participants.

[insert Table 1]

Examples of µFA maps in an FTD variant carrier and non-carrier are shown in **Figure 1**. FTD variant carriers exhibited lower µFA in the insula than non-carriers with large effect size: *F*(1,19)=5.89, 95% CI [-10.7, -0.75], *p*=0.027, *η*^*2*^*p*=0.26 **(Figure 2)**. The main effect of age showed a non-significant trend toward lower insula µFA: *F*(1,19)=3.83, *p*=0.067, η^*2*^*p*=0.18 **(Figure 3)**. No significant differences were observed between carriers and non-carriers for insular MD and volume (*p*>0.05, respectively). Greater insular MD and smaller insular volume were observed in older ages: *F*(1,19)=5.15, *p*=0.037, *η*^*2*^*p*=0.23, and *F*(1,19)=4.67, *p*=0.045, η^*2*^ *p*=0.22, respectively.

**Figure 1.**
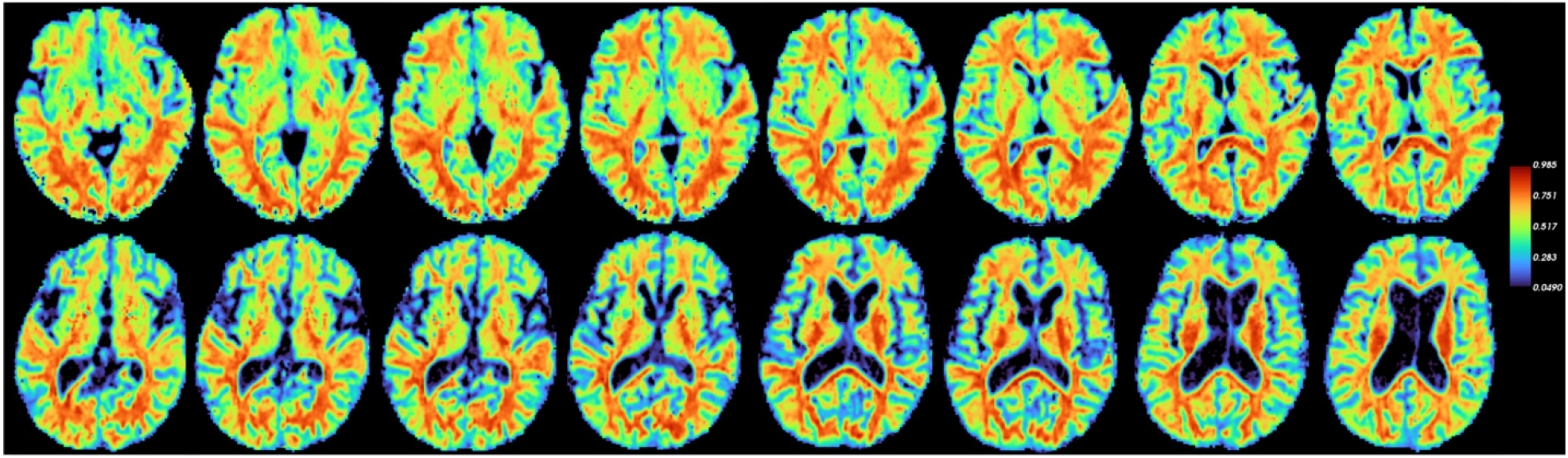
µFA maps for an FTD variant carrier (top panel) and non-carrier (bottom panel). Axial slices are in neurological orientation.

**Figure 2.**
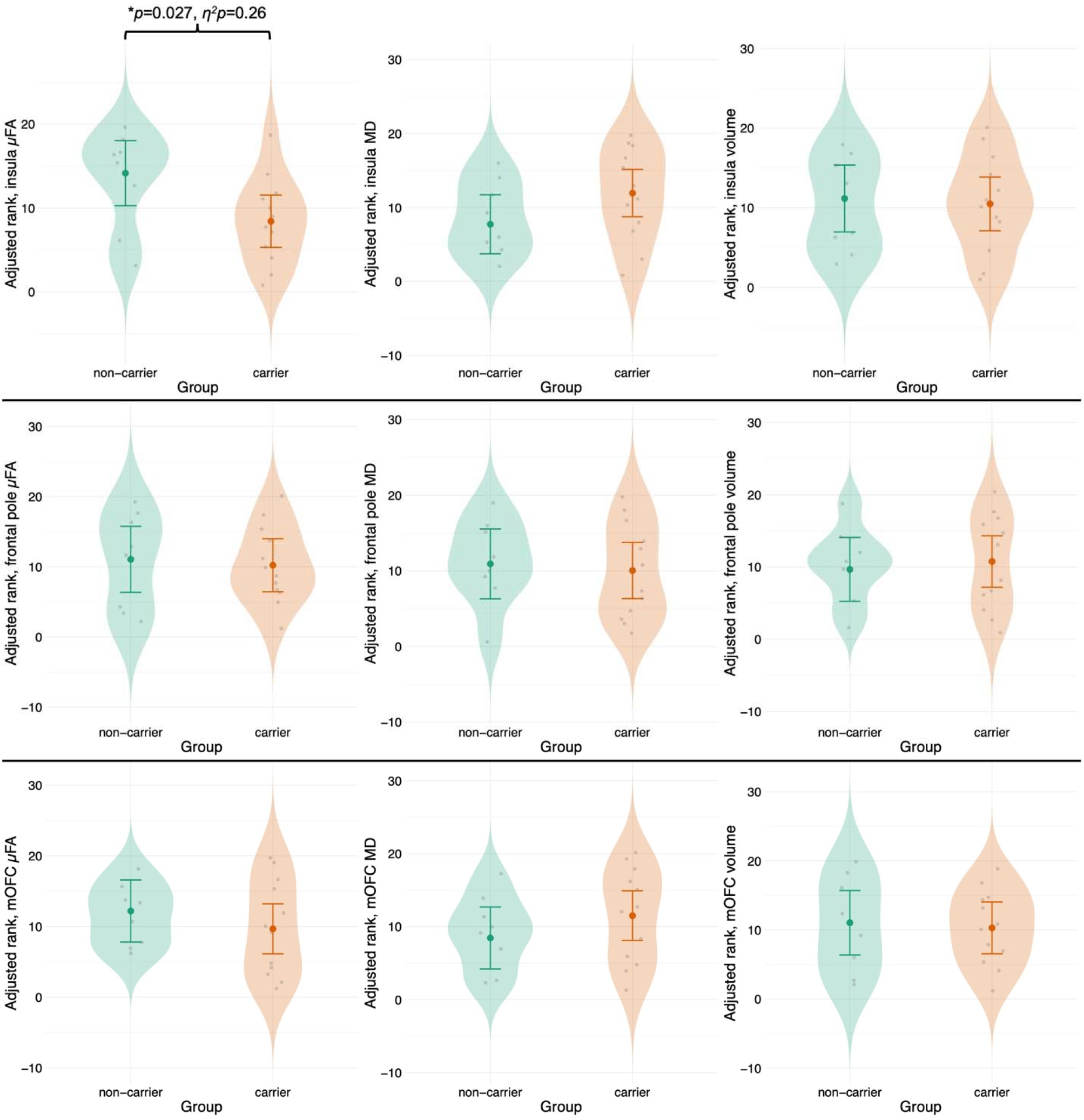
Average µFA, MD, and volumes of the insula (top panel), frontal pole (middle panel), and mOFC (bottom panel) between FTD variant carriers and non-carriers while adjusting for age. Lower µFA in the insula was observed in FTD variant carriers compared to non-carriers, without differences in MD and volumes. No differences in µFA were observed in the frontal pole or mOFC. \**p*<0.05. mOFC=medial orbitofrontal cortex.

**Figure 3.**
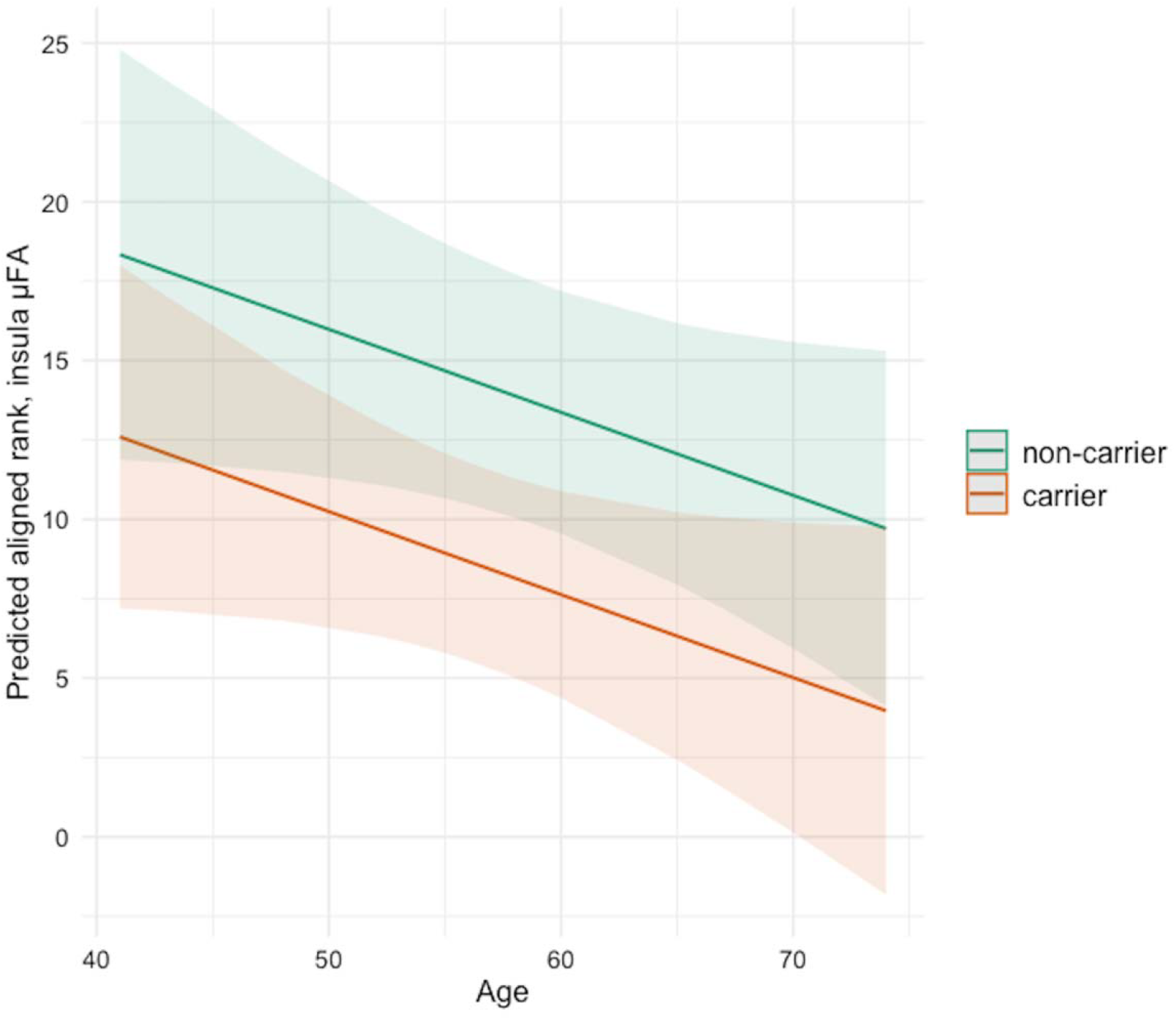
Predicted insular µFA is lower in carriers compared to non-carriers across age.

[insert Figures 1, 2, and 3 around here]

The post-hoc model for the insula was significant (*F*(3,18)=3.43, p=0.039, *η*^*2*^*p*=0.33), where, relative to non-carriers, reduced insula µFA was observed in symptomatic carriers, whose average age was 59 years (mean±SE: 5.88±2.24; *t*(18)=-2.57, *p*=0.019, 95% CI [-14.5, -1.98]). Mean µFA in pre-symptomatic carriers (mean±SE: 9.93±1.92; average age of 55 years) did not reach significance in comparison to non-carriers (mean±SE: 13.89±1.78; *t*(18)=-1.20, *p*=0.245, 95% CI [-9.58, 1.66]). Neither symptomatic nor pre-symptomatic carriers showed group differences in volume or MD (*p*>0.05 for all) **(Figure 4)**.

**Figure 4.**
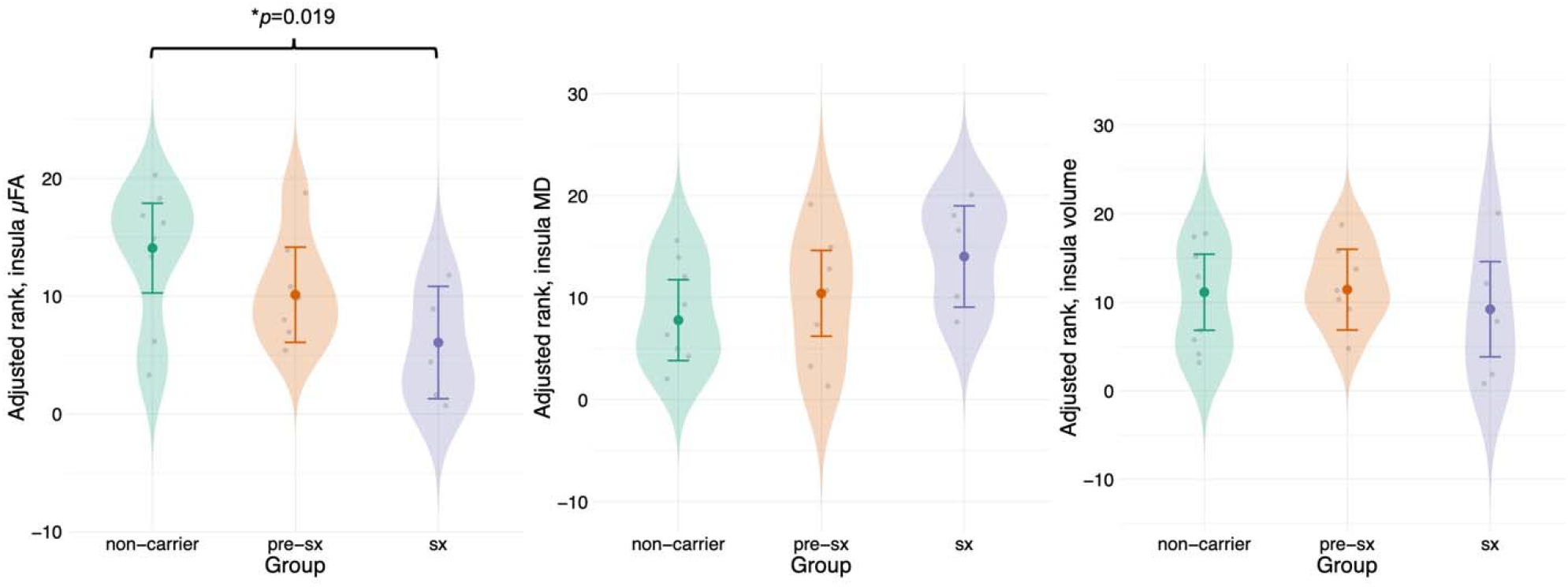
Reduced insula µFA was observed in symptomatic carriers compared to non-carriers.

[insert Figure 4]

Carriers and non-carriers did not differ in frontal pole µFA, MD, or volume (*p*>0.05). No significant effects of age were observed in the frontal pole µFA and volume models (*p*>0.05). Older participants trended towards having greater MD in the frontal pole: F(1,19)=3.44, *p*=0.081, η ^*2*^*p*=0.17.

Carriers and non-carriers also did not differ in µFA, MD, or volume of the mOFC (*p*>0.05). No main effects of age observed in any of these three models (*p*>0.05).

## DISCUSSION

This pilot study was the first to investigate µFA in genetic FTD. Reduced µFA of the insula was observed in FTD variant carriers compared to non-carriers, even when controlling for age, without differences in insula MD and volume. This aligns with established literature implicating the insula as a region that is selectively vulnerable to the earliest signs of neurodegeneration in FTD.^3,33–37^ Consistent with prior findings,^3,15,38^ this study observed age-related increases in insular MD and reductions in insular volume. Although greater age has been associated with reduced µFA in white matter tracts and subcortical regions of healthy adults,^18^ the between-group µFA difference observed in this study was more likely driven by genetic variant carrier status, and particularly by symptomatic participants, rather than the effects of age.

The insula, particularly the anterior subdivision, is associated with social and emotional processing and higher cognitive functions.^39^ This is because the anterior insula is reciprocally connected to the limbic cortex (including the anterior cingulate cortex, dorsolateral cortex, and amygdala),^40^ as well as to the orbitofrontal and anterior temporal areas.^39^ The dorsal anterior insula and frontoinsular regions are also part of the salience network, the aberrance of which has been suggested as a mechanism behind which anterior insula vulnerability to FTD-related neurodegeneration.^41,42^ Indeed, anterior insula atrophy has been implicated as a driver that initiates progressive widespread structural brain network atrophy in both genetic^33^ and sporadic^34^ forms of behavioural variant FTD. The anterior insula is also more prone to developing TDP-43 pathology than its posterior counterpart in multiple dementias, including FTD.^35^ Compared to healthy controls, altered resting-state functional connectivity^36^ and metabolic connectivity ^37^ between the anterior insula and FTD-associated regions have been reported in symptomatic behavioural variant FTD^36^ and asymptomatic *MAPT* P301L carriers.^37^ Reduced insula µFA reflects microstructure fibre loss, which could be due to loss of cortical gray matter neuronal axons and dendrites, or loss of glial process fibres, where the retraction of long astrocytic peri-synaptic processes^43^ and deramification of microglial processes^44^ occurs during “dying-back” patterns of reactive gliosis in neurodegeneration.^43–45^

Contrary to our hypotheses, we did not observe µFA differences in the frontal pole nor mOFC of FTD variant carriers compared to non-carriers, despite these regions typically being affected in FTD.^3,9,34,36^ The simplest explanation is that these cortical regions may not be as sensitive to gray matter microstructural losses at the carrier group’s median age of 55.5 years, with an even younger median age in pre-symptomatic participants. Event-based modelling suggested an early, focal trajectory of neurodegeneration beginning in the insula, followed by involvement of the mOFC and striatal regions, with subsequent extension to the medial frontal pole and temporal pole regions before spreading across the lateral frontal cortex.^9,34^ FTD brain charts were consistent with this temporal pattern of progressive neurodegeneration, with the anterior insula volumes in FTD diverging earliest from normal aging, followed by the medial frontal regions and then broader frontal cortex.^9^

There were several limitations to this pilot study. The sample size limited statistical power, preventing investigation of additional ROIs within *C9orf72-, MAPT-*, and *GRN*-associated FTD and robust comparison between presymptomatic carriers vs. non-carriers. This limited study interpretability and generalizability. Collection of larger samples in multi-centre studies is limited at the present time, as measuring µFA requires custom pulse sequences that are not yet available from vendors. While µFA can be sensitive to fiber content, other microstructural properties (e.g., cellular size, permeability, and exchange effects) may indirectly influence the parameter. Notably, to varying degrees, all diffusion MRI parameters are limited in the ability to attribute parameter differences to specific cellular components.

In conclusion, reduced insula µFA in genetic FTD variant carriers was observed compared to non-carriers, despite lack of difference in MD and volumes in this region, supporting that µFA may be more sensitive to granular microstructural integrity differences not detectable with conventional diffusion or T1-weighted MRI metrics. Replication in a larger sample with ROIs further tailored to gene specific patterns is warranted. Including µFA in multimodal analyses which combine structural and functional neuroimaging, neuropsychological testing, and early biomarkers of neurodegeneration, would likewise be helpful in assessing the potential utility of µFA as a biomarker of early neurodegeneration, especially given the increasing promise of clinical trials in genetic FTD.

## Data Availability

All data produced in the present study are available upon reasonable request to the authors.

## ACKNOWLEDGEMENTS

We thank all participants and their families for their time, effort, and contributions to this study.

